# A comparative analysis between a SIRD compartmental model and the Richards growth model

**DOI:** 10.1101/2020.08.04.20168120

**Authors:** Antônio M. S. Macêdo, Arthur A. Brum, Gerson C. Duarte-Filho, Francisco A. G. Almeida, Raydonal Ospina, Giovani L. Vasconcelos

**Affiliations:** Departamento de Física, Universidade Federal de Pernambuco, 50670-901, Recife, Brazil; Departamento de Física, Universidade Federal de Sergipe, 49100-000 São Cristóvão, Brazil; Departamento de Estatística, CASTLab, Universidade Federal de Pernambuco, 50740-540 Recife, Brazil; Departamento de Física, Universidade Federal do Paraná, 81531-990 Curitiba, Brazil

**Keywords:** COVID-19, Fatality curve, SIRD model, Richards growth model, Intervention strategies

## Abstract

We propose a compartmental SIRD model with time-dependent parameters that can be used to give epidemiological interpretations to the phenomenological parameters of the Richards growth model. We illustrate the use of the map between these two models by fitting the fatality curves of the COVID-19 epidemic data in Italy, Germany, Sweden, Netherlands, Cuba, and Japan.

## 1. Background

The pandemic of the novel coronavirus disease (COVID-19) has created a major worldwide sanitary crisis [1, 2]. Developing a proper understanding of the dynamics of the COVID-19 epidemic curves is an ongoing challenge. In modeling epidemics, in general, compartmental models [3] have been to some extent the tool of choice. However, in the particular case of the COVID-19 epidemic, standard compartmental models, such as SIR, SEIR, and SIRD, have so far failed to produce a good description of the empirical data, despite a great amount of intensive work [4, 5, 6, 7, 8, 9, 10, 11]. In this context, phenomenological growth models have met with some success, particularly in the description of cumulative death curves [12, 13, 14]. The recent discovery, within the context of a generalized growth model known as the beta logistic model [15], of a slow, power-law approach towards the plateau in the final stage of the epidemic curves is another remarkable example of this qualitative success. Growth models, however, have the drawback that their parameters may not be easily interpreted in terms of standard epidemiological concepts [16], as can the parameters of the usual compartmental models.

As a concrete example, consider the transmission rate parameter *β* of the SIR model [3]. It can be easily interpreted as the probability that a contact between a susceptible individual and an infective one leads to a transmission of the pathogen, times the number of contacts per day. Although the value of *β* cannot be measured directly in a model independent way, and it is probably not even constant in the COVID-19 epidemic curves, the epidemiological meaning of the parameter is nonetheless easy to grasp conceptually. As a result, models that incorporate such parameters in their basic equations are sometimes regarded as “more epidemiological,” so to speak, than others that do not use similar parameters. This state of affairs creates a somewhat paradoxical scenario, in which we have, on the one hand, the striking empirical success of phenomenological growth models sometimes being downplayed, owing to the lack of a simple epidemiological picture of the underlying mechanism [16], and, on the other hand, the failure of traditional epidemiological compartmental models to produce good quantitative agreement with the empirical COVID-19 data. A glaring instance of the inadequacy of standard compartmental models for the COVID-19 epidemic is their inability to predict the power-law behavior often seen in the early-growth regime as well as in the saturation phase of the accumulated death curves—a feature that is well captured by growth models [15], as already mentioned.

It is clear that a kind of compromise is highly desirable, in which we get the benefits of the accuracy of the growth models in describing the epidemic, along with a reasonable epidemiological interpretation of their free parameters. An attempt in this direction was presented by Wang [16], where an approximate map between the Richards growth model [16] and the accumulated number of cases of a SIR model was proposed. The two free parameters of the Richards model were expressed as a function of the epidemiological based parameters of the SIR model. Here we improve on this analysis in two ways: (i) we extend the SIR model to a SIRD model by incorporating the deceased compartment, which is then used as the basis for the map onto the Richards model; (ii) the parameters of the SIRD model are allowed to have a time dependence, which is crucial to gain some efficacy in describing realistic cumulative epidemic curves of COVID-19.

## 2. Data

It is in general very hard to estimate the actual number of infected people within a given population, simply because a large proportion of infections go undetected. This happens largely because many carriers of the coronavirus are either asymptomatic or develop only mild symptoms, which in turn makes the number of confirmed cases for COVID-19 a poor proxy for the actual number of infections. This issue is well known in the literature and referred to as the “under-reporting problem” [17, 18]. With this in mind, and in the absence of more reliable estimates for the number of infected cases, we shall here focus our analysis on the fatality curves, defined as the cumulative number of deaths as a function of time.

In the present study we considered the mortality data of COVID-19 from the following countries: Italy, Germany, Sweden, Netherlands, Cuba, and Japan. The data used here were obtained from the database made publicly available by the Johns Hopkins University [19], which lists in automated fashion the number of the confirmed cases and deaths attributed to COVID-19 per country. We have used data up to July 30, 2020.

## 3. Methods

### 3.1. The Richards Growth Model

The time evolution of the number of cases/deaths in an epidemy can be modelled by means of the Richards model (RM), defined by the following ordinary differential equation [20, 21, 22]:

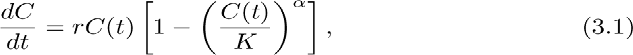

where *C*(*t*) is the cumulative number of cases/deaths at time *t, r* is the growth rate at the early stage, *K* is the final epidemic size, and the parameter *α* measures the asymmetry with respect to the s-shaped curve of the standard logistic model, which is recovered for *α*
 = 1. In the present paper we shall apply the RM to the fatality curves of COVID-19, so that *C*(*t*) will always represent the cumulative numbers of deaths at time *t*, where *t* will be counted in days from the first death.

Equation (3.1) must be supplemented with a boundary condition, which can be either the initial time, *t* = 0, or the inflection point, *t* = *t*_*c*_, defined by the condition *C*^*′′*^(*t*_*c*_) = 0, where *C*^*′′*^(*t*) = *d*^2^*C*(*t*)*/dt*^2^. A direct integration of (3.1) yields the following explicit formula:

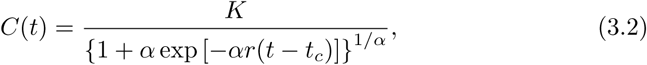

which will be the basis of our analysis.

### 3.2. SIRD model with constant parameters

We start by recalling the standard Susceptible (*S*)-Infected (*I*)-Recovered (*R*)-Deceased (*D*) epidemiological model

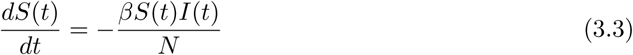

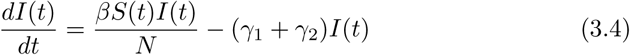

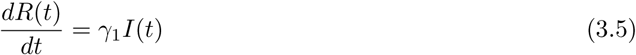

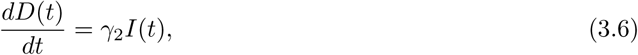

where *S*(*t*), *I*(*t*), *R*(*t*), and *D*(*t*) are the number of individuals at time *t* in the classes of susceptible, infected, recovered, and deceased respectively; whereas *N* is the total number of individuals in the population. i.e., *N* = *S*(*t*)+*I*(*t*)+*R*(*t*)+*D*(*t*). The initial values are chosen to be *S*(0) = *s*_0_, *I*(0) = *i*_0_, with *s*_0_ + *i*_0_ = *N*, and *R*(0) = 0 = *D*(0). The parameters *γ*_1_ and *γ*_2_ are the rates at which infected individual becomes recovered or deceased, respectively.

We then consider the following modified SIRD model, where in (3.3) and (3.4) we replace *N* with only the partial population in the *S* and *I* compartments, which takes into account the fact that the recovered (assuming they become immune) and the deceased cannot contribute to the transmission. We thus find

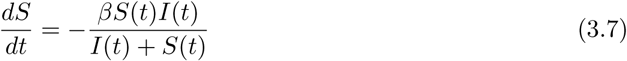

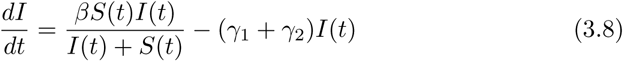

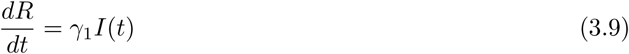

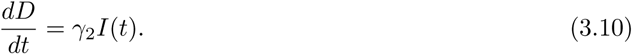

A fundamental quantity in epidemiology is the basic reproductive ratio, *R*_0_, which is defined as the expected number of secondary infections caused by an infected individual during the period she (or he) is infectious in a population consisting solely of susceptible individuals. In this model, *R*_0_ can be calculated using the next generation method [23, 24] and is given by

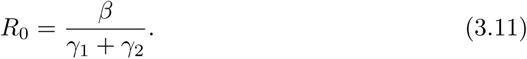

Next, we define *y*(*t*) = *S*(*t*) + *I*(*t*) and divide (3.8) by (3.7) to obtain

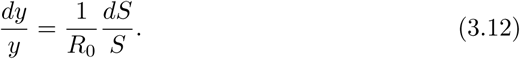

Integrating both sides of (3.12), and inserting the result into (3.7), yields a growth equation of the Richards type:

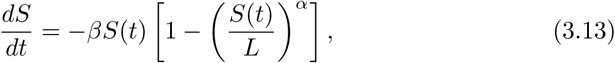

where *α* = 1 #x2212; 1*/R*_0_ and 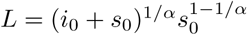.

We now seek to approximate the curve of accumulated death *D*(*t*), obtained from the SIRD model, with the Richards function *C*(*t*), as defined in (3.2). To this end, we first impose the boundary conditions *K* = *D*(*∞*) and *t*_*c*_ = *t*_*i*_, where *t*_*i*_ is the inflection point of *D*(*t*). By definition 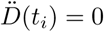, which implies from (3.10) that *İ*(*t*_*i*_) = 0 and thus

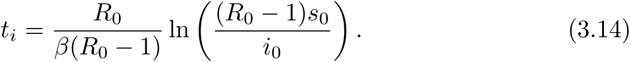

Furthermore, we require that at *t* = *t*_*i*_ both *C*(*t*) and its derivative *Ċ* (*t*) respectively match *D*(*t*) and 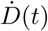, thus

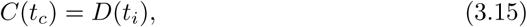

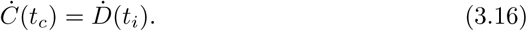

Using the condition *İ*(*t*_*i*_) = 0 in the SIRD equations, we find

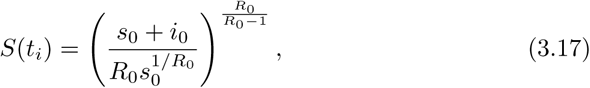

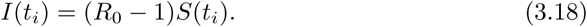

Using equations (3.15) and (3.16), we finally obtain the connection between the parameters (*r, α*) of the RM and the parameters (*β, γ*_1_, *γ*_2_) of the SIRD model

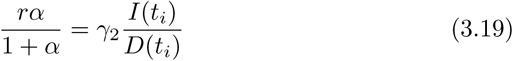

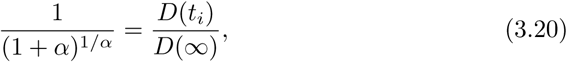

which are the central equations of this paper.

We can estimate the precision of the above ‘map’ between the RM and the SIRD model via the relative error function:

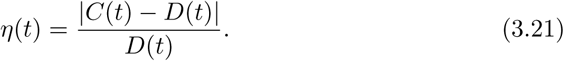

We have verified numerically that

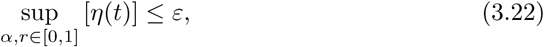

where *ε* is typically of order 0.1. A typical example of the agreement between the SIRD model, for a given set of parameters (*β, γ*1, *γ*2), and the RM with the parameters obtained from the map described by (3.19) and (3.20), is illustrated in Fig. 1. In Fig. 2 we show the simple monotonic dependence of the Richards parameters (*r, α*) on the parameter *β* of the SIRD model, for the biologically relevant interval 0≤*r, α ≤*1. We also show, for comparison, the behavior of the basic reproduction number *R*_0_.

**Figura 1:**
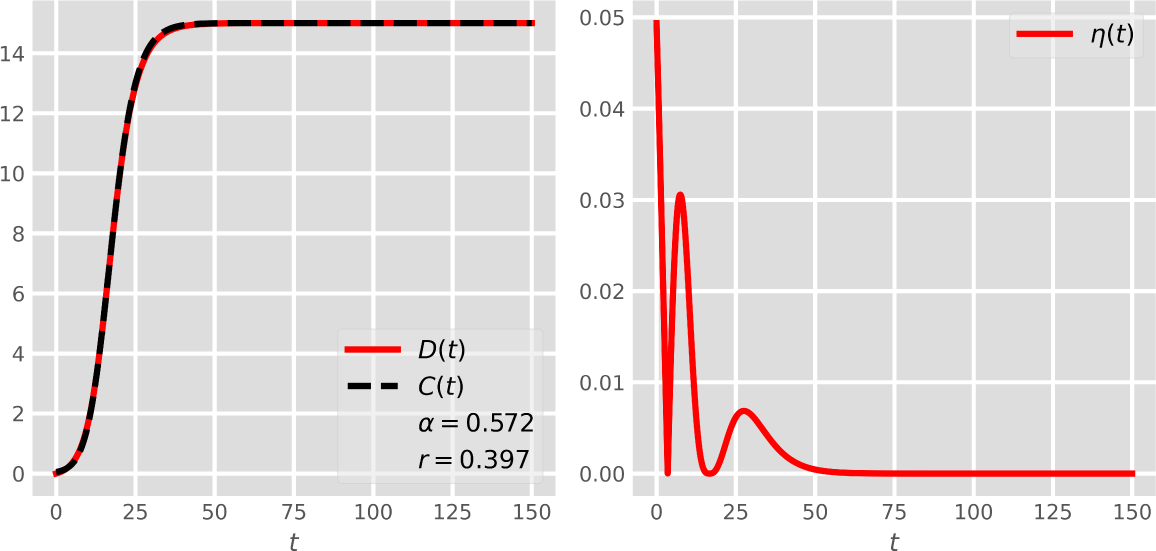
Illustrative picture (left panel) of the map between the Richards model and the SIRD model with constant parameters. The relative error function between the two models is shown in the right panel.

**Figura 2:**
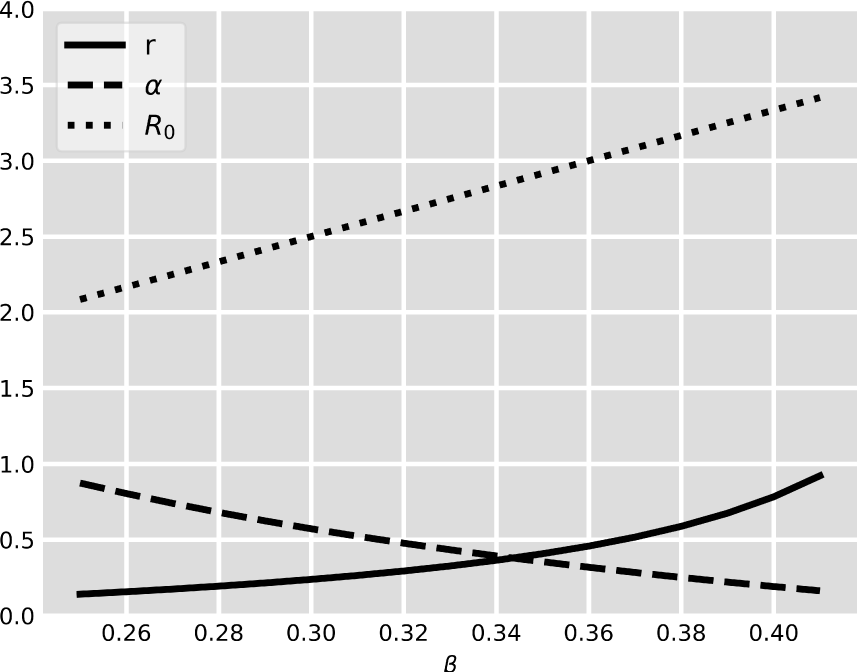
Behavior of the Richards parameters (*r, α*) and *R*_0_ as a function of the parameter *β* of the SIRD model.

### 3.3. SIRD model with time-dependent parameters

The SIRD model with constant parameters proved to be insufficient to accommodate properly the human intervention biased dynamics of the COVID-19 epidemics. The simplest solution to this problem is to allow the epidemiological parameter *β* to change in time according to the simple exponential decay function [4]

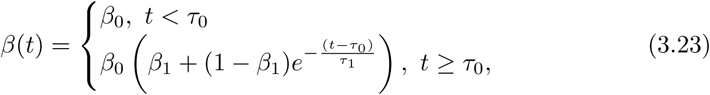

where *τ*_0_ is the starting time of the intervention and *τ*1 is the average duration of interventions. Here *β*_0_ is the initial transmission rate of the pathogen and the product *β*_0_*β*_1_ represents the transmission rate at the end of the epidemic. Remarkably, the central map equations, (3.19) and (3.20), are still valid, although *t*_*i*_ is no longer given by (3.14) and should be determined from the maximum of the curve *I*(*t*) obtained from the numerical solution of the SIRD equations, with the parameter *β* replaced by the function *β*(*t*).

## 4. Applications and Discussion

In Fig. 3 we demonstrate some applications of the SIRD-RM map by showing the cumulative number of deaths (red circles) attributed to COVID-19 for the following countries: Italy, Germany, Netherlands, Sweden, Japan and Cuba. In all figures shown, the continuous (black) curve is the numerical fit to the empirical data, as produced by the SIRD model with the time-dependent parameter *β*(*t*) given in (3.23), and the dashed (bright green) curve is the corresponding theoretical curve predicted by the RM, with the parameters as obtained from the map (3.19) and (3.20). The statistical fits were performed using the Levenberg-Marquardt algorithm [25], as implemented by the `lmfit Python` package [26], to solve the corresponding non-linear least square optimization problem. In other words, the *lmfit* package was applied to each empirical dataset to determine the parameters (*β*_0_, *β*_1_, *γ*1, *γ*_2_, *τ*_0_, *τ*_1_) of the SIRD model described in Secs. 3.2 and 3.3.

**Figura 3:**
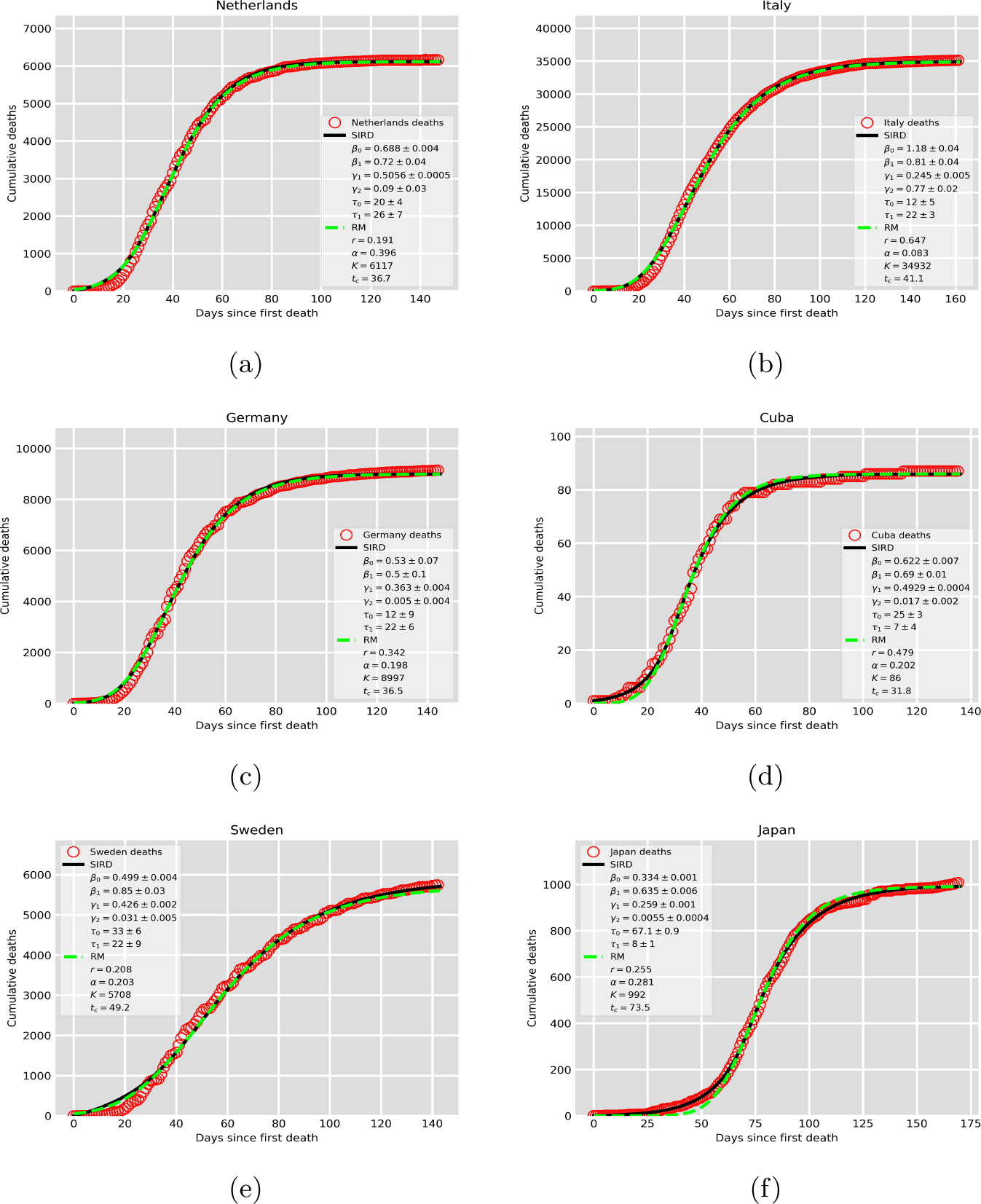
Cumulative number of deaths (red circles) attributed to COVID-19, up to July 30, 2020, for (a) Netherlands, (b) Italy, (c) Germany, (d) Cuba, (e) Sweden, and (f) Japan. The solid black curves are the best fits by the SIRD model with a time-dependent *β*(*t*), where the parameter estimates are given in the inset. The bright green dashes curve is the theoretical curve obtained from the Richards model, with the parameters computed from the numerically determined parameters of the SIRD model via the map (3.19) and (3.20).

One can see from Fig. 3 that the agreement between the RM and the SIRD model is very good in all cases considered, which satisfactorily validates the map between these two models. This result thus shows, quite convincingly, that the parameters of the Richards model do bear a direct relationship to epidemiological parameters, as represented, say, in compartmental models of the SIRD type. Although the interpretation of the Richards parameters (*r, α*) are less obvious, in that they involve a nonlinear relation with the probability rates used in compartmental models, these parameters should nonetheless be regarded as bonafide epidemiological parameters. Furthermore, it is important to emphasize the flexibility of the RM: this model, which has only two time-independent parameters, is equivalent (in the sense of the map discussed above) to a SIRD model with *time dependent* parameters. In other words, the two constant parameters of the RM are sufficient to characterize, to a rather good extent, the entire evolution of the COVID-19 epidemic in a given location.

It is worth pointing out that the discovery of power-law behaviors in the early-growth regime as well as in the saturation phase of the accumulated death curves, both of which are well described by the beta logistic model (BLM) [15], brings about the challenge to accommodate power laws into a compartmental model. A preliminary analysis [15] shows that substantial modifications in the SIRD equations may be required to achieve power law-behavior in the short-time and long-time regimes of the epidemic curves. The possibility of a map between the BLM and a modified SIRD model with time-dependent parameters is currently under investigation.

## 5. Conclusion

The present paper provides a map between a SIRD model with time dependent parameters and the Richards growth model. We illustrated the use of this map by fitting the fatality curves of the COVID-19 epidemics data for Italy, Germany, Sweden, Netherlands, Cuba and Japan. The results presented here are relevant in that they showcase the fact that phenomenological growth models, such as the Richards model, are valid epidemiological models not only because they can successfully describe the empirical data but also because they capture, in an effective way, the underlying dynamics of an infectious disease. In this sense, the free parameters of growth models acquire a biological meaning to the extent that they can be put in correspondence (albeit not a simple one) with parameters of compartmental model, which have a more direct epidemiological interpretation.

## Data Availability

The data for countries as well as for states/cities in the US are fetched from the COVID-19 Open Data Repository by CSSE/Johns Hopkins University (https://systems.jhu.edu

## Acknowledgements

This work was partially supported by the National Council for Scientific and Technological Development (CNPq) in Brazil, through the grants Nos. 303772/2017-4 (GLV), 312612/2019-2 (AMSM), and 305305/2019-0 (RO). AAB also thanks CNPq for its support through a PhD Fellowship (grant No. 167348/2018-3). RO thanks D.A.D.O. and CASTLab. The authors have declared no competing interest.

